# Scoping Review of Methods and Annotated Datasets Used to Predict Gender and Age of Twitter Users

**DOI:** 10.1101/2022.12.06.22283170

**Authors:** Karen O’Connor, Su Golder, Davy Weissenbacher, Ari Klein, Arjun Magge, Graciela Gonzalez-Hernandez

## Abstract

Real World Data (RWD) has been identified as a key information source in health and social science research. An important, and readily available source of RWD is social media. Identifying the gender and age of the authors of social media posts is necessary for assessing the representativeness of the sample by these key demographics and enables researchers to study subgroups and disparities. However, deciphering the age and gender of social media users can be challenging. We present a scoping review of the literature and summarize the automated methods used to predict age and gender of Twitter users. We used a systematic search method to identify relevant literature, of which 74 met our inclusion criteria. We found that although methods to extract age and gender evolved over time to utilize deep neural networks, many still relied on more traditional machine learning methods. Gender prediction has achieved higher reported performance, while prediction of age performance lags, particularly for more granular age groups. However, the heterogeneous nature of the studies and the lack of consistent performance measures made it impossible to quantitively synthesize results. We found evidence that data bias is a prevalent problem and discuss suggestions to minimize it for future studies.

## 1. Introduction

Real World Data (RWD)^1^ such as social media data has been increasingly recognized as a valuable resource for gaining knowledge and insight for a variety of health-related research topics including disease surveillance ^2,3^, pharmacovigilance ^4,5^, mental health ^6,7^. It can also be used for the identification of cohorts for potential recruitment into traditional studies ^8,9^. In short, social media can readily provide abundant personal health information in real-time.

The use of data from social media platforms such as Twitter, however, presents some inherent limitations for health-related research in that certain demographic information is not explicitly available through the Application Program Interface (API) ^10^. Knowing the demographics of users included in a study, including their age or gender, is important in health research. This information can be incorporated in analyses to identify disparities across demographic groups, to ensure inclusion of underrepresented groups and to elicit insights into age and gender differences in disease presentation or treatment response ^11,12^. Furthermore, given that Twitter users tend to be younger than the general population ^13^, knowing the specific demographic makeup of a cohort allows the researcher to report or make necessary adjustments to account for such bias.

Predicting demographic data is complex and challenging. A user’s profile does not necessarily include such information, and researchers have used other features available in the data, such as names, the content of the tweets, or the individual’s network. In this study, we present a scoping review of methods published since 2017 for determining the age and/or gender of Twitter users. We choose to focus our review on studies that use Twitter as the terms of use for this platform are well understood by both users and researchers, it includes an API, and the data is abundant for health related research ^14^.

While studies to predict Twitter user’s gender began as early as 2011 ^13,15–18^ and detecting the age of Twitter user’s has been addressed since 2013 ^19–21^, it is only since 2017 that the language processing community shifted its methods away from hand-crafted rules and represented the documents with dense vectors to train deep neural networks ^22,23^ resulting in a noticeable increase of performance for many applications. We sought to examine if these increases in performance were evident in the methods used for the prediction of the age and gender of Twitter users.

## 2. Methods

We conducted the study following the Preferred Reporting Items for Systematic and Meta Analysis extension for Scoping Reviews (PRISMA-ScR) ^24^ methodology. The PRISMA-ScR checklist is available in the Supplemental Information (SI) (Table S1). We searched several databases to identify research on the prediction of Twitter users’ age, gender, or both. We developed a detailed search strategy protocol. Our database search strategy combines three facets; facet one includes terms related to Twitter, facet two consists of terms for age or gender and facet three consists of terms for methods of prediction such as Machine Learning. The search strategy was translated as appropriate for each database. The detailed search strategy is available in the SI (Table S2). The machine learning term facet was expanded using terms from related reviews by Hinds and Joinson ^25^ and Abubakar ^26^. The search criteria were limited to peer-reviewed journals, conference proceedings, books, and theses.

The following databases were searched with a publication date range of 2017 or later (Figure 1).

**Figure 1:**
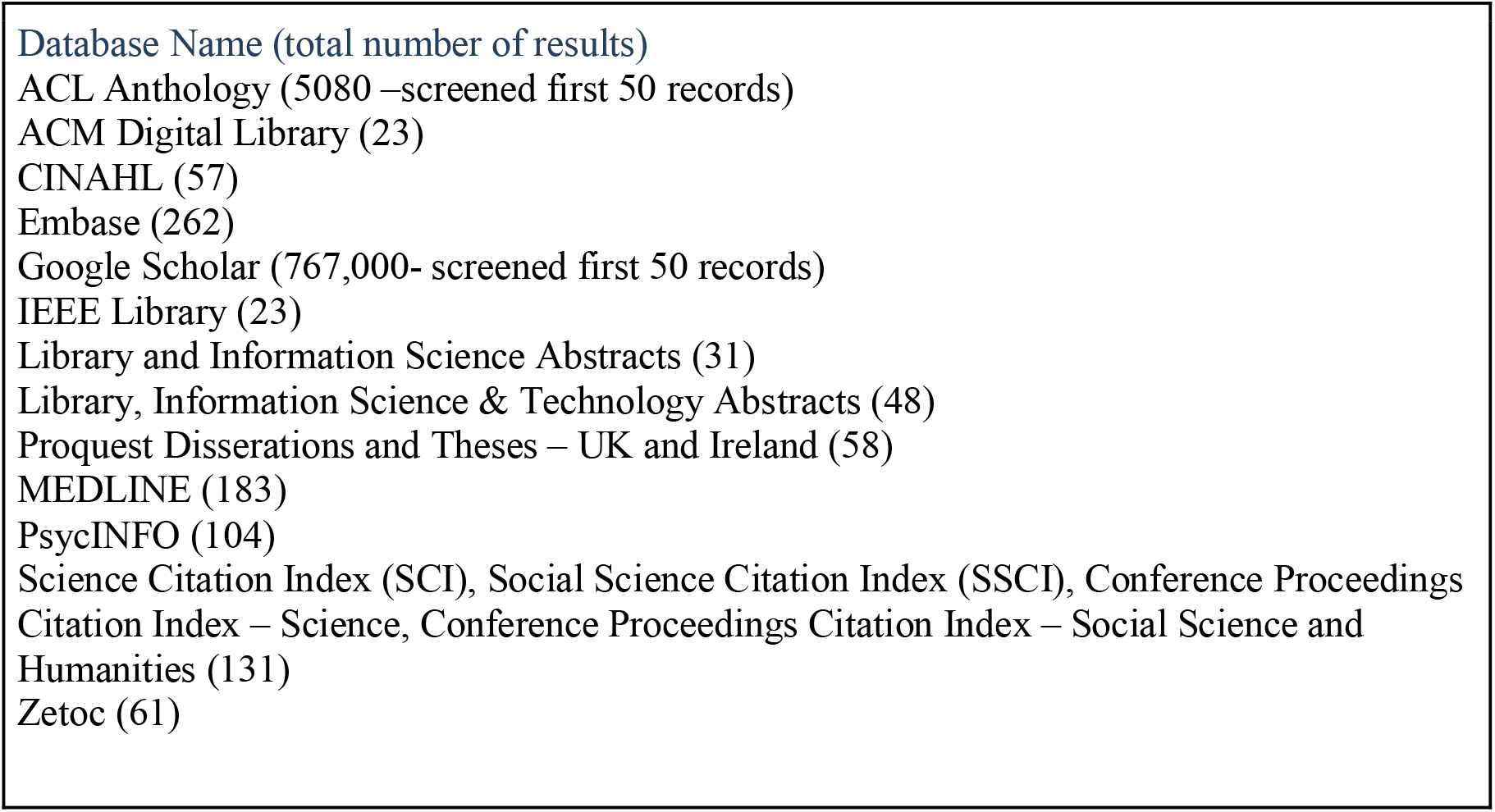
List of databases searched for reviews

Citations were exported to a shared Endnote library for deduplication. Using the PICOS ^27^ framework, we developed a list of inclusion and exclusion criteria (details below), and two screeners from the research team screened the results independently, with disputes on criteria discussed and a consensus decision reached. The first 50 records from both ACL and Google Scholar were screened separately using these same methods.

### 2.1 Inclusion and Exclusion Criteria

**Population-P**: Any Twitter data on Twitter users such as posts, profile details, photos or avatars. We excluded studies evaluating extraction from other types of social media.

**Intervention – I:** Methods to infer or predict gender or age demographic data of Twitter users. Articles that use machine learning, natural language processing, human in the loop or other computationally assisted methods to predict the gender or the age of users were included. Studies were excluded that contained no computation methods.

**Comparator – C:** Any or none.

**Outcome-O:** Gender or age prediction.

**Study Design - S:** Any type of peer-reviewed study reporting on the methods used to extract gender or age. Such information must be the primary focus of the study or reported in enough detail to be reproducible. Discussion papers, commentaries, and letters were excluded.

For reasons outlined in the introduction, we restricted the date of our search to only include publication from 2017 and beyond. No language restrictions were applied to the inclusion criteria; however, financial and logistical restraints did not enable translation from all languages.

### 2.2 Data Extraction

For each included article we extracted the following data: the year of publication, publication type (journal, conference paper, thesis), demographic extracted or predicted (gender, age, or both), the language of tweets, the size of the dataset, the collection method for the dataset, details of prediction models, features used in models (posts, profile, pictures), the performance of said models, the name of any software used for extraction, the measures used to evaluate methods and results of any evaluation, and the availability of data and/or code. The included articles were distributed amongst the authors to extract the data. The extracted data was validated by another author (KO).

### 2.3 Quality Assessment

This is a scoping review; thus no quality assessment is required.

### 2.4 Data Analysis

We summarized the performance stated in the papers. However, it is impossible to directly compare the approaches as the reported training data, validation methods, and performance metrics varied.

## 3. Results

Our database searches resulted in 981 records which were retrieved and entered into an Endnote library, where duplicates were removed, leaving 684 records for sifting.

From the abstract review, 172 references were deemed potentially relevant by either one of the independent sifters (SG and KO). The full-text of these articles was screened independently and disagreements discussed, resulting in 74 ^28–101^ that met our inclusion criteria and 98 excluded (Figure 2).

**Figure 2:**
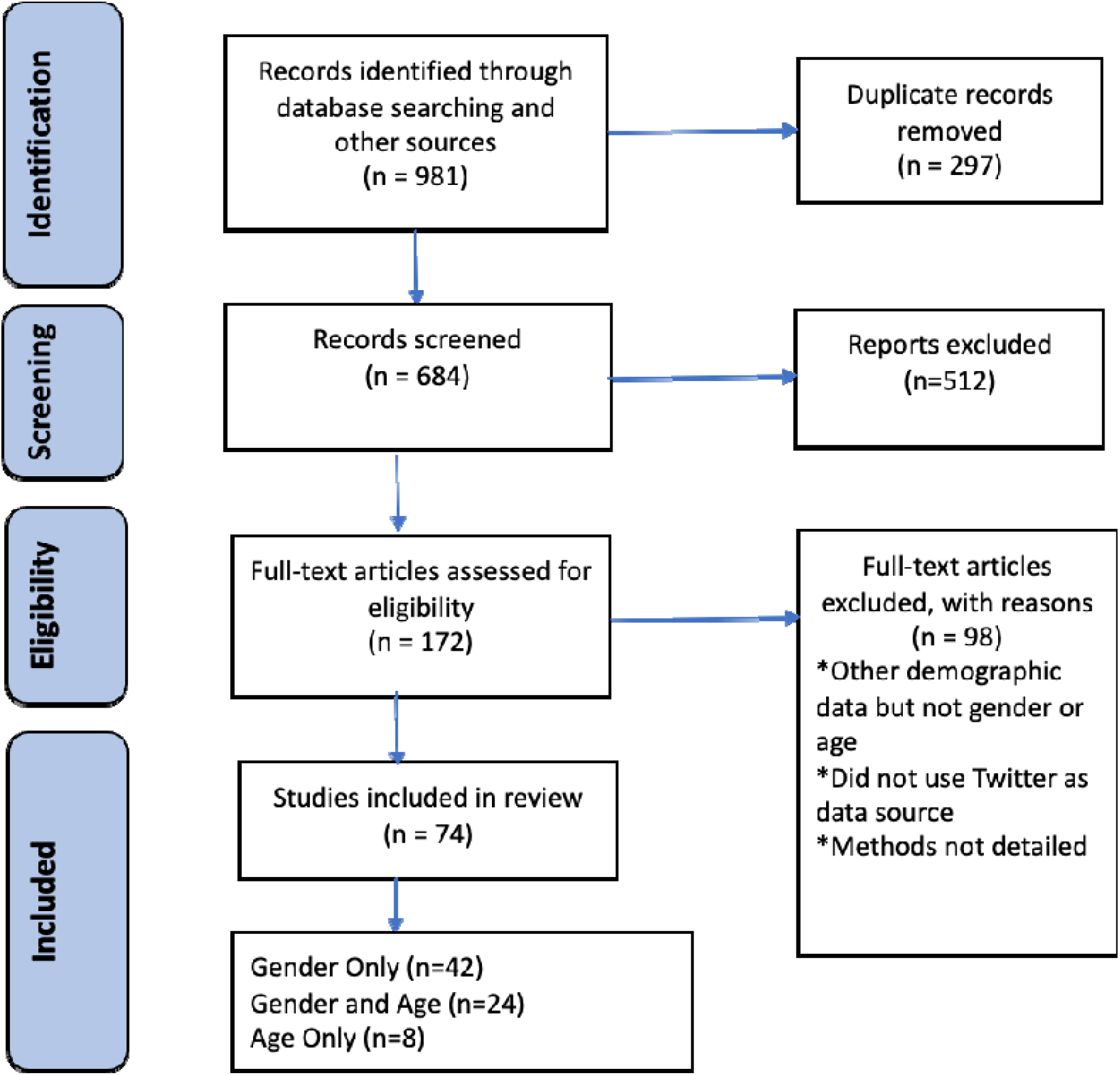
PRISMA flow diagram of included studies.

### 3.1 Characteristics of Included Studies

In the 74 studies (SI Table S3 and S4), the majority (n=42, 57.3%) focused on predicting the gender of the individual, 24 (32.0%) explored predicting both gender and age, and 8 (10.7%) focused solely on predicting age. Most of the studies were published in conference proceedings (n = 44, 58.7%), followed by journal articles (n = 28, 37.3%), thesis (n = 2, 2.7%) and in a book chapter (n = 1, 1.3%).

In 42 studies, developing methods to infer or predict the age and/or gender of Twitter users was the primary purpose of the study. In the remaining papers (n = 32), the identification of demographic characteristics of Twitter users was secondary. Within this last group, 9 studies developed ad hoc methods to determine age and/or gender, while the others used open-source models (n = 13) or off-the-shelf software (n=10).

### 3.2 Studies developing methods for gender and age prediction

#### 3.2.1 Gender

44 studies developed ad-hoc methods to predict the Twitter user’s gender. Of these, 32 predicted only gender ^28,29,31,33,36,37,47,48,50,51,54,55,58,60,61,64,65,68,71,72,75,81,83–86,90,92,94,96,100,101^ and gender was predicted along with the user’s age in 12 ^34,44,49,59,62,66,80,80,87,89,91,95^.

Most studies approached the problem of gender prediction as a binary classification task, predicting for an account the labels male or female, while three ^72,91,98^ added the classification of organization/brand.

We found that approaches to predict gender covered multiple languages, including English ^31,61,62,71,72,94,96^, German ^55^, Slovenian ^85^, Italian ^28^, Japanese ^68^, Arabic/Egyptian ^36,37,58^, French, Dutch, Portuguese, Spanish and a multilingual study including 28 languages/dialects ^91^.

##### 3.2.1.1 Datasets

For the training and validation of the approaches for gender detection, some studies used previously created annotated corpora, while others collected data directly from Twitter. Among the 19 studies that used previously annotated data sets, nine ^34,36,37,47,49,65,66,75,100^ used corpora from the PAN-CLEF author profiling tasks^102–108^, while ten studies ^51,54,62,64,72,83,84,94,96,101^ relied on data sets from other studies ^92,109–115^.

For studies that collected their own data, different components of the Twitter accounts were used. These components were used either for the purpose of manually or semi-automatically validating the gender of a user or for the purpose of computing features describing the user to train a classifier (SI Table S5). Despite data limitations from the Twitter API, it was the main source of data collection, with 18 studies ^28,30,31,33,48,50,51,55,58,62,68,71,85–87,95,96,100^ collecting data either as a random sample from the Twitter Streaming API or based on keywords or geographic location from the Twitter Search API. One study ^61^ collected data using a scraping tool, three ^59,91,92^ used a random sample from a collection of 10% of tweets from 2014-2017 or the Twitter archive, and one did not specify its data source ^44^.

The studies that created a labeled dataset (SI Table S6) to train and test, or to validate the performance of the system determined the gender of the users using multiple components of their Twitter accounts (SI Table S5). Eleven studies labeled the data through manual annotation where the annotators determined the gender using profile pictures ^31,33^, user names ^50^, profiles ^68^, or a combination of these ^55,61,71,85,87,89,95^. There were 11 studies that automatically, or (semi)-automatically, labeled their data sets through the detection of self-reports or gender identifying terms (e.g., mother, son, uncle, etc.) ^48,59,87,89,91,96^, the user’s name ^28,86,92^ or declarations on other linked social media ^95,96^. While three studies created their labeled datasets by using the accounts of famous social media influencers^57^ or other unspecified collection of users whose gender is known^28,43^. Of the 24 studies, only 8 reported data availability with most ‘by request’, only 2 have working links to the whole corpus (SI Table S6).

##### 3.2.1.2 Non-personal accounts

A Twitter account may not be authored by, or represent, a single person. There are organization or company accounts, as well as *bot* accounts. A bot is an automatic, or semi-automatic user account. Some bot accounts identify themselves as such and may be used to automatically amplify news or tweets related to a certain topics. Others may emulate human accounts and may be used with more malicious intent to sow discord, manipulate public opinion or spread misinformation. There were nine of the included studies ^28,55,71,72,75,82,83,85,91^ that removed non-personal organization accounts when they manually annotated their collections. Other studies implemented heuristics to explicitly detect and remove non-personal accounts ^28,29,38,50,60,86,92,101^, bots ^77^ or both ^58,116^. Others used previously annotated dataset either consisting of only personal accounts, or labeled non-personal accounts which were removed, or collected their datasets based on self-reports. The remaining studies provided no details of how, or if, these accounts were removed (SI Table S5).

##### 3.2.1.3 Features and Models

The data labeled with user’s gender was used in the reviewed studies to build and evaluate classification models based on features describing the text in tweets (e.g., n-grams, word embeddings, hashtags, URLs) ^36,37,44,47–50,54,58,61,65,66,71,75,83,88,92,100^, or in the users’ profile metadata (e.g., user names, bio, followers, users followed) ^28,30,31,51,59,64,91,94,101^, a combination of their profile metadata and tweets ^31,33,55,62,72,86,87,89,96^, images ^31,59,87,91,95^. There was one study from Japan that included the user’s geographic information under the assumption that, culturally, a person of a certain demographic is more likely to frequent specific places ^68^.

Among the systems using hand crafted features (n = 25, 56.8%), most achieved their best results using Support vector machine (SVM) ^28,33,44,51,61,64,65,83–85,92,95,117^, while others utilized logistic regression ^66,86,89^, Naïve Bayes ^30,71^ random forests ^59^, bag of trees ^49^, XGBoosting ^68^, an ensemble of three supervised approaches ^58^ (Table 1). Others used deep learning methods (n = 15, 34.1%) such as deep neural networks (DNN), convolutional neural networks (CNNs), feed forward neural networks (FFNN) or recurrent neural networks (RNN) ^34,47,50,54,72,94,100^, BiLSTM ^37^, gated recurrent units (GRU) ^36^, graph recursive neural networks (GRNN) ^62^ and multi-modal deep learning networks ^87,91^. One study created a meta-classifier ensemble classifying users based on the predictions of multiple individual classifiers ^96^, including SVM, BERT and two existing models ^91,118^. Another created a deep neural network for learning with label proportion (LLP), a semi-supervised approach ^31^. Results for the best-performing deep learning model as reported in each study are included in Table 2. Studies that employed lexical matching (n=4, 9.1%) of the user’s name to a curated names dictionary ^29,60,80,81^ to determine gender reported no validation or performance metrics.

**Table 1:** Top reported system performance for studies inferring gender of Twitter users using traditional ML methods. Result metrics are reflected here as reported in the original publications and are not comparable to each other.

| First Author (year) | Language<sup>a</sup> | ML Method<sup>b</sup> | Reported Performance |  |
| --- | --- | --- | --- | --- |
|  |  |  | F1-score | Accuracy |
| Cesare (2017) | en | ensemble: lexical match/SVM/DT | 0.84 | 0.83 |
| Jurgens (2017) | en | RF ensemble | 0.78 | 0.8 |
| Ljubešić (2017) | pt/fr/nl/es/de/it | SVM | NA | 0.61 - 0.69 |
| Markov (2017) | en/es/nl/it | LogR | NA | 0.57 - 0.77 |
| Mukherjee (2017) | en | NB | 0.75 | 0.71 |
| Verhoeven (2017) | sl | SVM | 0.93 | 0.93 |
| Volkova (2017) | en/es | LogR | NA | 0.82 |
| Xiang (2017) | en | SVM/PME | NA | 0.76 |
| Cheng (2018) | en/fi/Taglish | SVC with Lasso | 0.84 | 0.84 |
| Emmery (2018) | en | Fasttext | NA | 0.76 |
| Giannakopoulos (2018) | NA | SVM PNN | NA | 0.87 |
| Khandelwal (2018) | Code-mixed hi-en | SVM | NA | 0.9 |
| Miura (2018) | ja | XGBoosting | NA | 0.89 |
| Van der Goot (2018) | en/nl/fr/pt/es | SVM | NA | 0.66 - 0.72 |
| Alessandra (2019) | it | ensemble: lexical match/SVM | NA | 0.75 |
| Hirt (2019) | de | ensemble: binary classifiers | 0.81 | NA |
| Hussein (2019) | Dialect eg ara | ensemble: RF/LinR | NA | 0.77 - 0.88 |
| Vicente (2019) | en/pt | ensemble: face ++/LinR/SVM | NA | 0.93 - 0.97 |
| Arafat (2020) | in | Multinomial NB | NA | 0.75 |
| Baxevanakis (2020) | gr | SVM | NA | 0.7 |
| Garcia-Guzman (2020) | en | Bag of Trees | 0.64 | 0.64 |
| López-Monroy (2020) | en/es | Bag of Trees | 0.64 | 0.64 |
| Pizarro. (2020) | en/es | SVM | 0.82 - 0.84 | NA |
| Vashisth (2020) | en | LogR | NA | 0.57 |
| Wong (2020) | en | SVM | 0.58-0.62 | 0.60 |

<sup>a</sup> ara: Arabic, de: German, eg: Egyptian, en: English, es: Spanish, fr: French, gr: Greek, hi: Hindi; it: Italian, in: Indonesian, ja: Japanese, nl: Dutch, pt :Portuguese, sl: Slovenian
<sup>b</sup> DT: decision trees, LDA: latent Dirichlet allocation, LinR: linear regression, LogR: logistic regression, mm: multi-modal, NB: naive Bayes, RF: random forest, SVM: support vector machine

**Table 2:** Top reported system performance for studies inferring gender of Twitter users using deep learning ML methods. Result metrics are reflected here as reported in the original publications and are not comparable to each other.

| First Author (year) | Language<sup>a</sup> | ML Method<sup>b</sup> | Reported Performance |  |
| --- | --- | --- | --- | --- |
|  |  |  | F1-score | Accuracy |
| Ardehaly (2017) | en | DeepLLP | 0.96 | NA |
| Geng (2017) | en | ensemble: LDA/CNN | NA | 0.87 |
| Kim (2017) | en | GRNN | NA | 0.68 |
| Vijayaraghavan (2017) | en | DMT | 0.89 | NA |
| Wang (2017) | NA | CNN | 0.91 | 0.9 |
| Bayot (2018) | en/es | CNN | NA | 0.59 -0.72 |
| Bsir (2018) | ara | GRU | NA | 0.79 |
| Wood-Doughty (2018) | en | RNN | 0.84 | 0.84 |
| Bsir (2019) | ara | BILSTM with Attention | NA | 0.82 |
| Hashempour (2019) | pt/fr/nl/es/de/it | FFNN | NA | 0.84 - 0.86 |
| Wang (2019) | multilingual | mmDNN | 0.92 | NA |
| ElSayed (2020) | eg/ara dialects | multichannel CNN-biGRU | NA | 0.84 - 0.91 |
| Oyasor (2020) | en | DNN | NA | 0.68 |
| Zhao (2020) | en | CNN | 0.80 | NA |
| Yang (2021) | en | ensemble: M3/SVM | 0.95 | 0.94 |

<sup>a</sup> ara: Arabic, de: German, eg: Egyptian, en: English, es: Spanish, fr: French, it: Italian, nl: Dutch, pt
<sup>b</sup> BiLSTM: bidirectional long-term short-term memory, CNN: convolutional neural network, DMT: deep multi-modal multitask, DNN: deep neural network, GRNN: graph recurrent neural network, GRU: gated recurrent networks, LLP: learning with label proportions, mmDNN: multi-modal deep neural network, RNN: recurrent neural network

##### 3.2.1.4 Performance

Performance results from traditional machine learning against deep learning methods cannot be meaningfully compared as they are evaluated against different corpora, including the different languages, their size, as well as non-standardized reporting metrics. However, looking at the overall results in terms of F1-score, the results of studies using deep learning had a relatively narrower range of reported performance (0.84 – 0.96), with a higher minimum of 0.84 and higher maxiumum of 0.96, compared to the reported performance range for traditional ML methods, which spans from 0.64 to 0.93.

#### 3.2.2 Age

We found in those that developed ad hoc methods, 19 studies that sought to predict the Twitter user’s age, where 7 predicted only age ^32,43,45,52,69,73,74^. All but one of the studies ^59^ approached the detection of Twitter users’ age as automatic classification of predefined age groups. The number of age groups varies across the studies (Table 3), with the ages categorized into two ^32,52,62,89,95^, three ^30,45,69,73,74,80,87^, four ^49,91^ or more ^34,43,44,66^ groups. The range of ages within the groups also varies across the studies; for example, across the five studies that take a binary classification approach, Guimaraes et al., ^52^ use *13-19* and *20+* as the two age groups, Volkova et al ^89^ and Kim et al, ^62^ used 18-23 or 25+, Xiang et al., ^95^ used 30 or below or above 30, while Ardehaly and Culotta ^32^ use *less than 25* and *25+*. Except for two studies that did not report the language of the tweets used ^30,52^, all studies used English language tweets. Eight of the studies extended their systems to include additional languages including Spanish ^34,43,66,89^, Dutch ^66,73,74^, Filipino ^44^, and multiple languages ^91^.

**Table 3:** Top reported system performance for studies inferring age of Twitter users using traditional ML methods. Result metrics are reflected here as reported in the original publications and are not comparable to each other. Reviews are ordered by number of classification groups.

| First Author (year) | Number of Age Groups | Age Class Detail | Language <sup>a</sup> | ML Method<sup>b</sup> | Reported Performance |  |
| --- | --- | --- | --- | --- | --- | --- |
|  |  |  |  |  | F1-score | Accuracy |
| Jurgens (2017) | NA | continuous | en | RF regression | NA | 0.71 |
| Volkova (2017) | 2 | 18-23, 25-30 | en/es | LogR | NA | 0.77 |
| Xiang (2017) | 2 | <=30, >30 | en | CPME | NA | 0.74 |
| Ardehaly (2018) | 2 | <25 and >25 | en | LLP | NA | 0.78 |
| Morgan-Lopez (2017) | 3 | 13-17, 18-24, >24 | en | LogR | 0.74 | NA |
| Arafat (2020) | 3 | <=24, 25-39, >=40 | NR | LogR | NA | 0.71 |
| Cornelisse (2020) | 3 | 18-24, 25-54, >55 | en | LogR | 0.78 | NA |
| Markov (2017) | 5 | 18-24, 25-34, 35-49, 50-64, 65+ | en/es/nl/it | LogR | NA | 0.56 - 0.65 |
| Cheng (2018) | 5 | 18-24, 25-34, 35-44, 45-54, 55-64 | en/fi/Taglish | SVC | 0.61 | 0.86 |
| Garcia-Guzman (2020) | 4 | 18-24, 25-34, 35-49, 50+ | en | Bag of Trees | NA | 0.67 |
| Chamberlain (2017) | 10 (3) | <12, 12-13, 14-15, 16-17, 18-24, 25-34, 35-44, 45-54, 55-64, >64 | en/es/fr/pt | Bayesian Probability | 0.31<br>0.86 (3 Class) | NA |

<sup>a</sup> en: English, es: Spanish, pt :Portuguese, fr: French, nl: Dutch, de: German, it: Italian, in: Indonesian, fi: Filipino, NR: not reported
<sup>b</sup> LLP: learning with label proportions, LinR: linear regression, LogR: logistic regression, NB: naive Bayes, RF: random forest, SVM: support vector machine

##### 3.2.2.1 Datasets

While most of the studies that developed new algorithms prepared new data sets to evaluate the algorithms with data retrieved directly using Twitter’s API ^30,32,45,52,69,87^ or using other sources or methods ^43,59,91^ (SI Table S4), several used data sets made available from others’ studies to train and/or evaluate their algorithms. Two ^73,74^ combined data sets from ^19,21,69^ Kim et al.,^62^ used the dataset from Volkova et al., ^119^, while three ^34,49,66^ used data sets that were created for the PAN-CLEF author profiling shared tasks ^103–105^. The studies that prepared new data sets (SI Table S6) labeled users’ age groups by (semi-)automatically searching for (1) tweets that self-report birthday announcements or age ^32,59,69,87,89,91^, (2) tweets in which a user is wished a happy birthday ^69^, (3) profiles that self-report age ^43,45,87,91^, (4) profiles that mention age-related keywords (e.g., *grandparent*) ^45,91^, (5) manual annotation based on images or profile metadata ^91,95,119^, or (6) by subjectively perceiving age groups based on the content of individual tweets ^52^. In one study ^30^, a mixture of self-reported data and individuals with known demographic information was used to label the data. Similar to the studies on gender, the reported availability of the corpora is scare. Only 5 studies reported that their datasets are available, 2 by request, 1 provided a link to the whole dataset and 2 proved link to a sample of the corpus (SI Table 6).

##### 3.2.2.2 Features and Models

The studies used the labeled age groups to evaluate classification models based on features of the users’ profile metadata (e.g., user names, bio, followers, users followed) ^30,32,43,59,91^, a combination of their profile metadata and tweets (e.g., n-grams, word embeddings, hashtags, URLs) ^52,62,69,73,74,87,89^, tweet texts only ^44,45,49,66^ or images ^59,87,91,95^.

For automatic classification, the majority of studies (12/19, 63.2%) used traditional supervised machine learning methods including logistic regression ^30,45,66,69,89^, Bayesian probabilistic inference ^43^, random forests ^59^, bag of trees ^49^, Support vector machine (SVM) ^44,95^ or a semi-supervised approach, learning from label proportion (LLP) ^32^. Others used deep learning methods (7/19, 36.8%) such as convolutional neural networks (CNNs) ^34,52,73,74^, graph recursive neural networks (GRNN) ^62^ and multi-modal deep learning networks ^87,91^. Results of the best performing systems for each study are reported in Tables 3 and 4. One study ^80^ classified age based on a previously developed age lexicon and did not report any performance metrics.

**Table 4:**
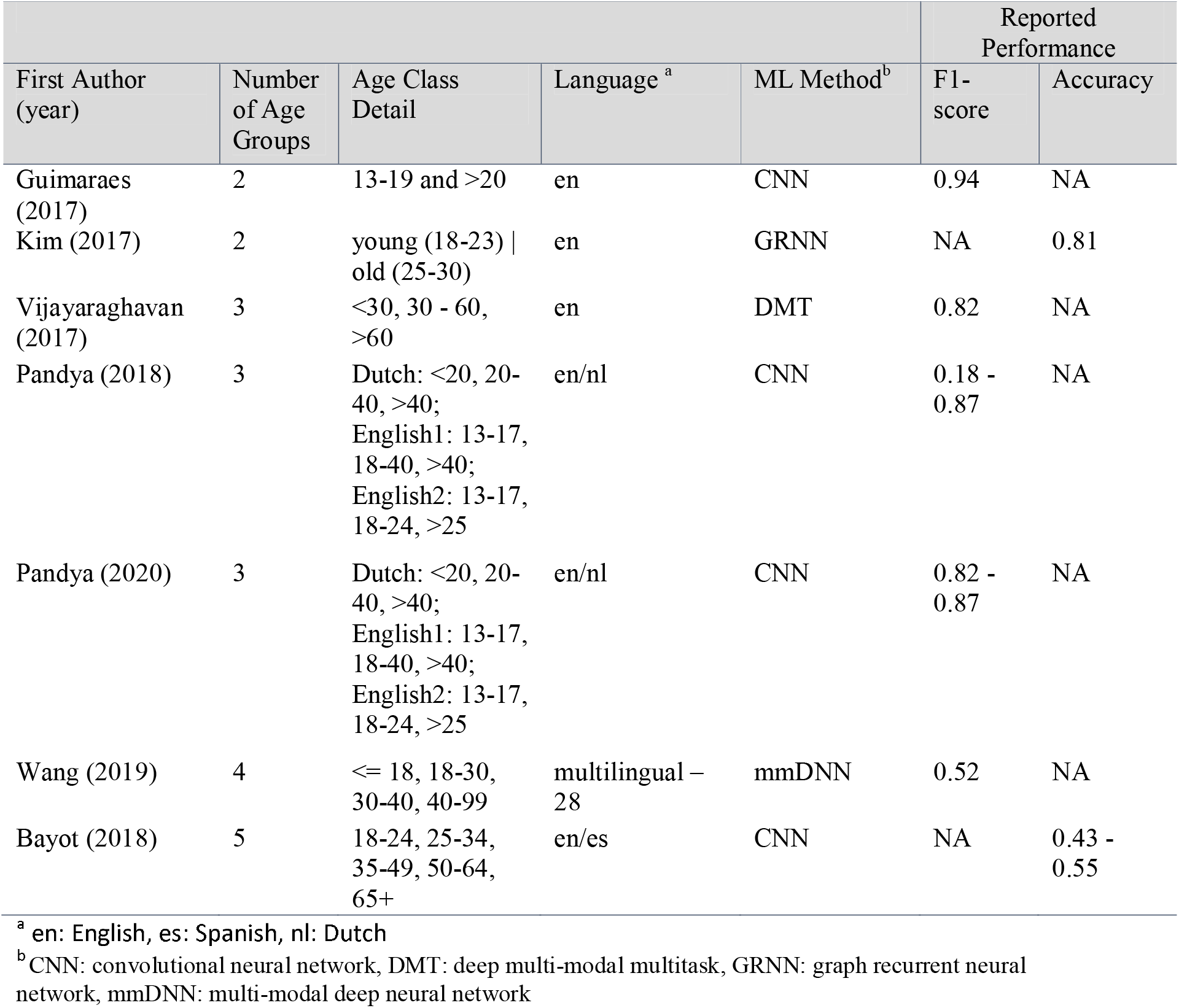
Top reported system performance for studies inferring age of Twitter users using deep learning ML methods. Result metrics are reflected here as reported in the original publications and are not comparable to each other. Reviews are ordered by number of classification groups.

##### 3.2.2.3 Performance

Given the variation in classification (e.g., different age groupings, different number of classification categories) and reported performance metrics it is difficult to assess performance difference between studies using tradition machine learning versus those using deep learning or neural networks. However, for binary and ternary classification, there is a slight improvement in classification performance over quaternary or more classification.

### 3.3 Studies using previously developed methods

Within our included studies, there were 23 for which the detection of gender or age was secondary to their research and previously developed methods were used to detect the demographic information for their cohort. Of the 23, 13 used open-source models, and ten used off-the-shelf software.

#### 3.3.1 Open-Source Models

Three of the studies ^53,78,79^ drew upon an extant model ^120^ that employs a predictive lexicon for multi-class classification of age groups or gender for their applications. None of these studies created a validation corpus to assess the performance of the system which was originally reported as 89.9% accuracy for gender and 0.84 Pearsons correlation coefficient for age. One study ^97^ utilized the same text-based model ^120^ and an image model ^121^ to find age and gender of their cohort. When tested against their gold standard corpus of self-reports from profile descriptions, they found the imaging model performed best for gender (accuracy = 90 – 92%), while textual features gave the best results for age (accuracy = 60%). Three studies ^57,70,93^ used Demographer ^118,122,123^ for gender predictions, with one ^70^ evaluating the performance against a set of users who had self-reported their gender in a survey finding an F1-score of 0.869 for women and 0.770 for men. Two studies ^40,41^ used their ensemble classifier of previously developed models, with a reported accuracy of 0.83 and F1-score of 0.83^101^.Two studies ^46,99^ used M3 ^91^ to detect gender and age, with one validating the performance using a manually labeled dataset finding for gender the performance achieved 95.9% accuracy and an F1 score of 0.957 and for age 77.6% accuracy and F1 score of 0.731. One^35^ used DEX ^124^ for age and gender detection which reported a validation error of 3.96 years for age and an 88% accuracy for gender. One study ^77^ used the rOPenSci “gender” package, no assessment of the performance was reported

#### 3.3.2 Off-the-shelf software

For the 10 studies that used off the shelf software, Face ++ was the most common, being used by 6 studies^42,56,67,76,88,98^. The remaining used DemographicsPro ^38,39^,Microsoft Face API ^63^ and RapidMiner ^82^.

In four of the studies ^67,76,82,88^ no validation of performance was carried out and a further two simply reported that DemographicsPro requires 95% confidence ^38,39^. Others compared to manual annotation and identified accuracy for age using Face ++ at 82.8% ^56^ or 68% for strict age groups or 83% if the age groupings were relaxed ^42^. The performance for age using Microsoft Face API was measured at 0.895 Gwet’s AC ^63^, when compared to manually labeled datasets.

For gender, those studies that measured accuracy using their own gold standard set of users accuracy was recorded as 94.4% ^56^ or 88% ^42^ using Face ++. Other studies ^67,76,88^ reported the confidence level reported by Face ++ for gender prediction of 95% +-0.015.

Only one study ^98^ went beyond manual annotation to create a gold standard and used multiple search techniques to manually verify age and sex, including LinkedIn profiles, electoral roll listings, personal websites, Twitter descriptions, and Twitter profile images. In it, Face++ accuracy for age was reported as 40.4% and sex 44.8% (with valid images age 32.5% and gender 87.7%) and crowdsourcing annotation accuracy for age was 60.8% and gender 86.4% (with valid images for age 56.1% and gender 93.9%).

## 4. Discussion

In this review, we aimed to provide an overview of recent machine learning methods being used to predict the gender and age of Twitter users. Our review indicates that the identification of gender has received more attention. However, despite the popularity of both tasks, no accepted standards for research (data collection and evaluation) has emerged, resulting in a large number of heterogeneous studies which are difficult to compare. In consequence, it is difficult to conclude where the state-of-the-art stands for these tasks. Data collection practices can severely bias the datasets, thus resulting in correspondingly biased automatic methods. We note below some recommendations to avoid the noted biases, summarized in Figure 3.

**Figure 3.**
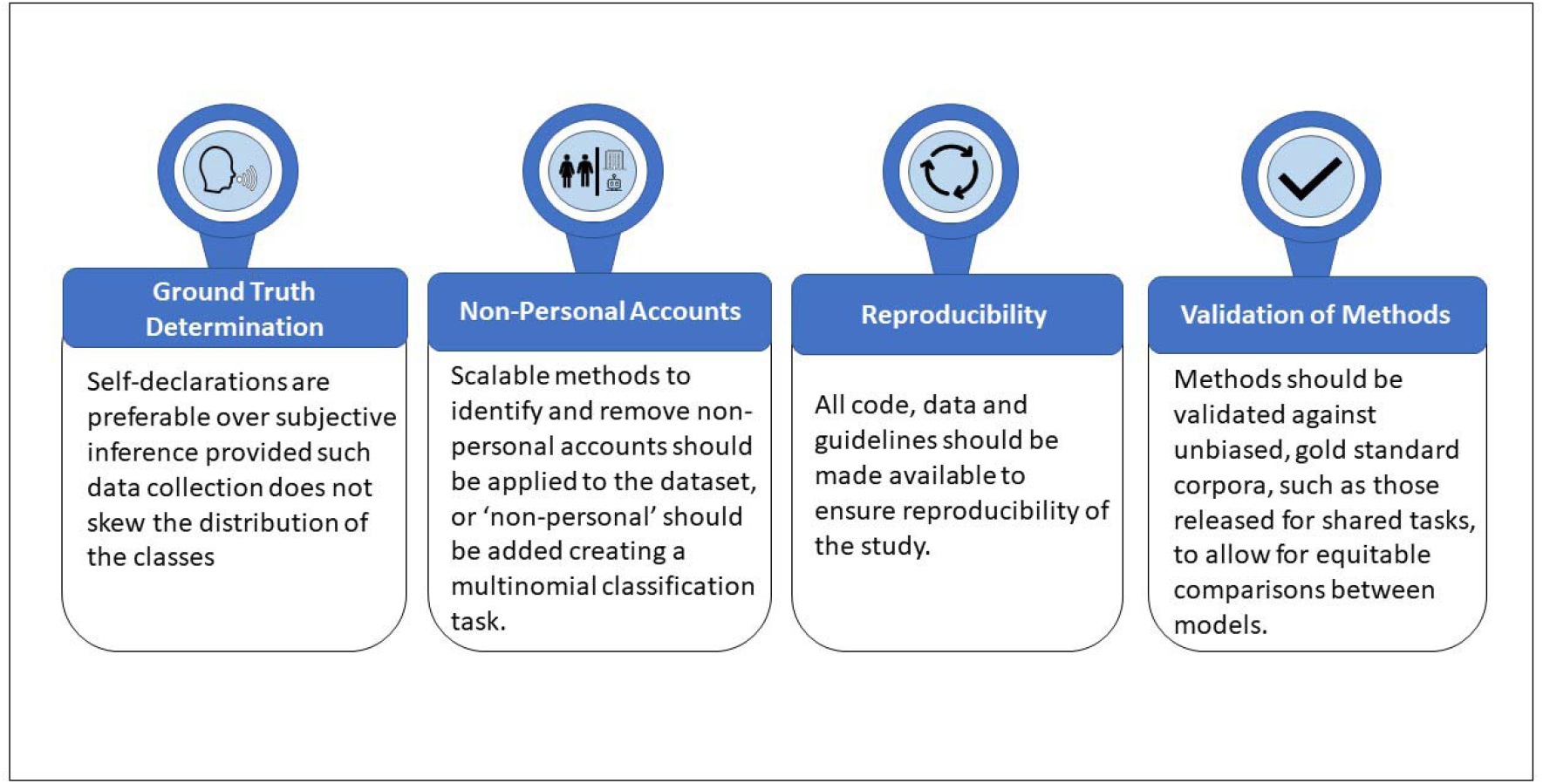
Summary of recommendations for best practice.

Demographic information is an important task to address in order to fully realize the potential advantages of using social media data such as that from Twitter in health-related research. In the US, the National Institute of Health (NIH) has committed to include women participants in clinical studies and to include sex as a biological variable, finding that the disaggregation of data by sex will allow for sex-based comparisons. A recent review ^125^ found this disaggregation in the development of machine learning models led to the discovery of sex-based differences that improved model performance for sex-specific cohorts. Age is also important as age can affect the course and progression of disease ^125^, or the effects of medication ^126^. Given the significance of this information, it is important that accurate and reproducible models be developed. One way to ensure the reproducibility of the models is for researchers to make available all data and code, including annotation guidelines. In addition to model performance, studies that create annotated corpora should report annotator agreement measures in order to assess the quality of the corpus. Few of the included studies made available their data or code (SI tables S2, S3 and S6).

A particular difficulty when comparing different systems comes from a lack of a ‘gold standard’ labeled corpus to compare the systems against. Some studies created their own corpora, collecting data randomly or based on keywords relevant to their studies. Others reused datasets from prior studies or from shared tasks. Although outside the scope of this review, there have been shared tasks which aim to advance research through competition, focusing on gender and age prediction. A longstanding shared task focused on author profiling has been hosted at the PAN workshop at the Conference and Labs of the Evaluation Forum (CLEF) ^102–108^. More recently, Social Media Mining for Health (SMM4H), 2022, included 2 tasks for age detection releasing new annotated corpora for the tasks ^127^, Several researchers reported utilizing the corpora from these shared tasks. Testing and reporting performance metrics against these publicly available data sets, without alteration, would provide a comparable metric of different approaches. However, while reusing annotated corpora provides quick access to labeled data, it does have some limitations, including data loss over time as users delete their Tweets, which not only reduces the size of the data but also can result in a data imbalance of the corpus.

### 4.1 Gender Prediction

Almost all included studies approached the gender prediction task as a binary classification task, identifying a user as either male or female. We note that, even when focusing on binary gender classification, which is the prevalent approach, the task of gender prediction on Twitter could be better characterized as a multinomial classification task: given a user account, the classifier should return male, female, or “non-personal”. The last label (non-personal) could account for Twitter users representing organizations or bots. While some studies attempted to identify and exclude non-personal accounts as a preprocessing step, others developed their systems using previously annotated datasets which were exclusively labeled as male or female users or removed non-personal accounts during annotation before training and testing. It is unknown how well these systems would perform when extended to unseen data which may contain non-personal accounts.

Excluding non-personal accounts, the ratio of males to females in the training dataset is also important, as it should mimic the natural distribution of Twitter users estimated to be 31.5% females and 68.5% males as of January 2021. However, some authors biased their collections by using unconventional methods of collection or using datasets that were artificially balanced. The most conventional method to collect a set of Twitter accounts is to query from the Twitter API any tweet mentioning functional words without semantic meaning such as “of”, “the”, or “and”. Whereas collecting Twitter users using functional or neutral keywords, a given language, or geographic areas, resulted in a male/female ratio close to the ratio naturally observed on Twitter, other choices resulted in collections with different ratios. Such change of ratios could have improved (or deteriorated) the training of the authors’ classifiers and biased their evaluations which were not reflecting the performance of their approach on a random sample of users taken from Twitter.

All studies treated gender as a binary determination of male or female. While some referenced the limitation of this approach, they opted to use these designations given the need to align their data with outside resources such as US census or social security administration data. We note that gender, unlike biological sex, it is not necessarily binary as a social construct and has been shown to influence a person’s use of healthcare, interactions, therapeutics response, disease perceptions and decision making ^128^, This underlies the importance of expanding the efforts of classification beyond binary to improve accuracy and avoid misinterpreting results.

### 4.2 Age Prediction

The prediction of age task generally had lower performance that gender prediction. This was true for studies that developed their own models as well as those using open source or off-the-shelf software. This may be because most approached it as a multiclass classification task. The proxies used, such as language, names, networks or images, may have limited predictive value for age. Additionally, the demographics of Twitter users means that any data set will be inherently imbalanced, providing few training examples for age groups in the tail ends of the distribution. This data imbalance may lead to too few instances of the minority classes to effectively train the classifier. For models that classified based on images, this poor performance for age may be unsurprising given that it can be difficult for humans to discern age from a single image. In addition, photos may be subject to photo editing or enhancing, or not be a recent photograph of the user. Due to a lack of error analysis reports in the included studies, it is difficult to determine the source of the classification difficulty for age.

Performance aside, the fact that the number and range of the age groups vary across the studies suggests that a classification approach is not generalizable to all research applications. Identifying exact age, rather than age groups, can generalize to applications that do not align with predefined groupings of binary or multi-class models; however, using high-precision rules to extract self-reports of exact age from user’s profile metadata had shown not to scale. As we worked on this study, we noted that none of the systems reviewed opted for extracting exact age. To test the feasibility and utility of a generalizable system that extracts exact age from tweet in a user’s timeline using deep learning methods, we developed a classification and extraction pipeline using the RoBERTa-Large model and a rule-based extraction model^129^. The system was trained and tested on 11000 annotated tweets. The classification of tweets mentioning an age achieved an F1-score of 0.93 The extraction of age from these tweets achieved a F1-score of 0.86. From a collection of 245, 947 users, an age was extracted for 54% using REPORTage. A shared task for the classification task was run at the SMM4H 2022 workshop, in which we released the annotated dataset. We did not include our approach in the scoping review as there were no comparable systems published before the release of the approach as part of the SMM4H 2022 shared task.

### 4.3 Potential bias of differing methods

Limitations of using names to distinguish between genders may promote bias, particularly if the names used for training do not represent the ethnic diversity of the population and some cultures may have more unisex names than others which cannot be used to distinguish gender. There can be a high degree of uncertainty for many users for which gender cannot be classified by name, estimates by Sloan 2013^130^ are that 52% will be unclassified using this method. However, studies have suggested that those classified may be relatively accurate given that data from UK Twitter demonstrates a high level of agreement with UK census data ^131^. Furthermore, used alone, this heuristic may label some organization accounts, such as PAUL_BAKERY, as a person but it removes most of the organizations’ accounts.

Relying on self-declarations may be prone to bias as well. For example, regarding age, younger people are more likely to profess their age than older adults as age may be more important to them and with respect to gender pronouns these may be more likely to be declared by those in particular occupations or of a particular social class. Indeed, there may also be other bias to self-declarations of data by culture, background, social class, or country of origin or residence.

Using users’ profile images is challenging for gender and age identification. Not all Twitter users provide a picture of themselves with many opting for pictures of their pets, objects, children, scenery or even celebrities. Even those with pictures of themselves can be problematic if the quality is poor, the picture contains more than one face, or the picture is not recent particularly for inferring age. A comparison of systems using images to infer demographics ^132^ not only measured the accuracy of identifying age and gender but also the share of images for when a face is detected finding only around 30% of Twitter users had a single detectable face.

Methods to filter out organizations in the studies included removing accounts with large number of followers ^50^, or searching explicitly for organization by looking up user name terms linked to economic activities such as restaurant, hotel etc. ^28^. These methods remove accounts that are not representing a single user, however, they do not remove bots. While one study created a classifier to detect bots, the filtering of bots was limited to those identified in manual annotation, by simple heuristics, or non-existent in many of the studies (SI Table S5).

### 4.4 Validation of age and gender proxies

For those cases where age or gender are estimated it is necessary to conduct validation exercises whereby the data are compared to a ‘gold standard data set’ to establish accuracy levels. For example, one study ^98^ that used off-the-shelf software also created a manually annotated gold standard dataset for measuring accuracy. This study found that although the accuracy of crowdsourcing was higher than software, it was only around 60% for age. This puts into question the use of manual annotations alone as a gold standard.

The most reliable way of generating a ‘gold standard’ is to obtain this information directly from the user. This may be in form of direct correspondence with the user such as messaging via social media, or the other way around: requesting Twitter handles in surveys that collect demographic data. Other methods for validation, such as manual extraction, may be less rigorous. However, these can be improved by multiple independent annotators, using experienced teams.

External validation of the model is also a vital step to assess how the model will perform on unseen data. In a validation on a second dataset, Yang et al, ^96^ found performance dropped on all but two of their models, stressing the importance of benchmarking existing systems on a targeted corpus. This step is equally important when using existing systems so a range of expected performance can be reported and used in any analysis of the output.

### 4.5 Limitations

It is unlikely that we have identified all studies using off-the-shelf software as we did not search for specific named software, but part of our remit was to identify the array of software used. We did not limit our inclusion to only studies that developed their own software, therefore, we have included studies that used proprietary software. These software products do not publish their methodologies; therefore, we are unable to directly compare these approaches to others.

We also included studies for which the extraction of age and gender was needed for the primary focus of their study. These studies either used proprietary software, previously developed methods or developed limited methods to infer the demographic information. In general, these studies did not report the performance of their inference methods on their datasets. While some reported the original performance metrics of the methods used, the assumption cannot be made that these methods will perform similarly across all data.

## 5. Conclusions

The extraction of demographic data such as age and gender is an important step in increasing the value and application of social media data. Many methods are reported in the literature with differing degrees of success. While we sought to explore whether deep learning approaches would advance the performance for these tasks as it has been shown to do for other NLP tasks, many of the included studies utilized traditional machine learning methods. Though only explored by a handful of studies, deep learning methods appear to perform well for the prediction of a user’s gender or age. However, direct comparison of methods is impossible at this point. This highlights the need for recently developed, publicly available gold-standard corpora, such as those released for shared tasks like SMM4H or PAN@CLEF, in order to have unbiased data and baseline metrics to compare different approaches against.

## Data Availability

The search strategy and extracted data on included studies is available in the Supplementary Data.

## Author Contributions

SG, KO and GG devised the study and identified data for extraction. SG created and executed the search strategy and created the initial draft of the manuscript. SG and KO were responsible for study selection. All authors were responsible for data extraction, summarization and discussion. KO synthesized all data and created all tables. All authors commented on and edited the manuscript. KO provided the final version of this manuscript.

All authors contributed to the final draft of the manuscript.

## Data availability

The search strategy and extracted data on included studies is available in the Supplementary Data.

