## Supplemental Tables 1,2 for "Scoping Review of Methods and Annotated Datasets Used to Predict Gender and Age of Twitter Users"

**Table S1: Preferred Reporting Items for Systematic reviews and Meta-Analyses extension for Scoping Reviews (PRISMA-ScR) Checklist**

| **SECTION** | **ITEM** | **PRISMA-ScR CHECKLIST ITEM** | **REPORTED ON PAGE #** |
| --- | --- | --- | --- |
| **TITLE** | | | |
| Title | 1 | Identify the report as a scoping review. | 1 |
| **ABSTRACT** | | | |
| Structured summary | 2 | Provide a structured summary that includes (as applicable): background, objectives, eligibility criteria, sources of evidence, charting methods, results, and conclusions that relate to the review questions and objectives. | 1 |
| **INTRODUCTION** | | | |
| Rationale | 3 | Describe the rationale for the review in the context of what is already known. Explain why the review questions/objectives lend themselves to a scoping review approach. | 1 - 2 |
| Objectives | 4 | Provide an explicit statement of the questions and objectives being addressed with reference to their key elements (e.g., population or participants, concepts, and context) or other relevant key elements used to conceptualize the review questions and/or objectives. | 1 - 2 |
| **METHODS** | | | |
| Protocol and registration | 5 | Indicate whether a review protocol exists; state if and where it can be accessed (e.g., a Web address); and if available, provide registration information, including the registration number. | NA |
| Eligibility criteria | 6 | Specify characteristics of the sources of evidence used as eligibility criteria (e.g., years considered, language, and publication status), and provide a rationale. | 3 |
| Information sources* | 7 | Describe all information sources in the search (e.g., databases with dates of coverage and contact with authors to identify additional sources), as well as the date the most recent search was executed. | 2 - 3  Table S2 |
| Search | 8 | Present the full electronic search strategy for at least 1 database, including any limits used, such that it could be repeated. | Table S2 |
| Selection of sources of evidence† | 9 | State the process for selecting sources of evidence (i.e., screening and eligibility) included in the scoping review. | 3 - 4 |
| Data charting process‡ | 10 | Describe the methods of charting data from the included sources of evidence (e.g., calibrated forms or forms that have been tested by the team before their use, and whether data charting was done independently or in duplicate) and any processes for obtaining and confirming data from investigators. | 3 |
| Data items | 11 | List and define all variables for which data were sought and any assumptions and simplifications made. | 3 |
| Critical appraisal of individual sources of evidence§ | 12 | If done, provide a rationale for conducting a critical appraisal of included sources of evidence; describe the methods used and how this information was used in any data synthesis (if appropriate). | NA |
| Synthesis of results | 13 | Describe the methods of handling and summarizing the data that were charted. | 3 - 4 |
| **RESULTS** | | | |
| Selection of sources of evidence | 14 | Give numbers of sources of evidence screened, assessed for eligibility, and included in the review, with reasons for exclusions at each stage, ideally using a flow diagram. | 3 - 4 |
| Characteristics of sources of evidence | 15 | For each source of evidence, present characteristics for which data were charted and provide the citations. | 3 - 11 |
| Critical appraisal within sources of evidence | 16 | If done, present data on critical appraisal of included sources of evidence (see item 12). | NA |
| Results of individual sources of evidence | 17 | For each included source of evidence, present the relevant data that were charted that relate to the review questions and objectives. | 3 - 11 |
| Synthesis of results | 18 | Summarize and/or present the charting results as they relate to the review questions and objectives. | 7 – 9, Tables 1 -4 |
| **DISCUSSION** | | | |
| Summary of evidence | 19 | Summarize the main results (including an overview of concepts, themes, and types of evidence available), link to the review questions and objectives, and consider the relevance to key groups. | 11 - 15 |
| Limitations | 20 | Discuss the limitations of the scoping review process. | 15 |
| Conclusions | 21 | Provide a general interpretation of the results with respect to the review questions and objectives, as well as potential implications and/or next steps. | 14 - 15 |
| **FUNDING** | | | |
| Funding | 22 | Describe sources of funding for the included sources of evidence, as well as sources of funding for the scoping review. Describe the role of the funders of the scoping review. | 16 |

*From:* Tricco AC, Lillie E, Zarin W, O'Brien KK, Colquhoun H, Levac D, et al. PRISMA Extension for Scoping Reviews (PRISMAScR): Checklist and Explanation. Ann Intern Med. 2018;169:467–473. [doi: 10.7326/M18-0850](http://annals.org/aim/fullarticle/2700389/prisma-extension-scoping-reviews-prisma-scr-checklist-explanation).

Table S2: Search strategies and results for individual databases.

| Database | Date Searched | Search Strategy | Results |
| --- | --- | --- | --- |
| ACL Anthology | 26/06/2020 | (Twitter OR tweet OR tweeting OR tweets OR retweet* OR tweep*) AND (gender OR age OR demograph*) | 5,080 records |
| ACM Digital Library | 26/06/2020 | [[Publication Title: twitter] OR [Publication Title: tweet] OR [Publication Title: tweeting] OR [Publication Title: tweets] OR [Publication Title: retweet*] OR [Publication Title: tweep*]] AND [[Publication Title: age] OR [Publication Title: gender] OR [Publication Title: demograph*] OR [Publication Title: minor*] OR [Publication Title: baby boomer*] OR [Publication Title: "generation x"] OR [Publication Title: "generation y"] OR [Publication Title: "generation z"] OR [Publication Title: "gen x"] OR [Publication Title: "gen y"] OR [Publication Title: “gen z$] OR [Publication Title: millennial*] OR [Publication Title: adult*] OR [Publication Title: teen*] OR [Publication Title: youth*] OR [Publication Title: adolescen*] OR [Publication Title: juvenile*] OR [Publication Title: young] OR [Publication Title: youngster*] OR [Publication Title: aged] OR [Publication Title: aging] OR [Publication Title: ageing] OR [Publication Title: elder*] OR [Publication Title: old] OR [Publication Title: retired] OR [Publication Title: older*] OR [Publication Title: geriatr*] OR [Publication Title: gerontolog*] OR [Publication Title: senior*] OR [Publication Title: senescen*] OR [Publication Title: retiree*] OR [Publication Title: sexagenarian*] OR [Publication Title: septuagenarian*] OR [Publication Title: octagenarian*] OR [Publication Title: nonagenarian*] OR [Publication Title: centenarian*] OR [Publication Title: supercentenarian*] OR [Publication Title: veteran*] OR [Publication Title: pensioner*] OR [Publication Title: male*] OR [Publication Title: female*] OR [Publication Title: men] OR [Publication Title: women]]  (Twitter OR tweet OR tweeting OR tweets OR retweet* OR tweep*) AND (age OR gender OR demograph* OR minor* OR baby boomer* OR "generation x" OR "generation y" OR "generation z" OR "gen x" OR "gen y" OR “gen z$ OR millennial* OR adult* OR teen* OR youth* OR adolescen* OR juvenile* OR young OR youngster* OR aged OR aging OR ageing OR elder* OR old OR retired OR older* OR geriatr* OR gerontolog* OR senior* OR senescen* OR retiree* OR sexagenarian* OR septuagenarian* OR octagenarian* OR nonagenarian* OR centenarian* OR supercentenarian* OR veteran* OR pensioner* OR male* OR female* OR men OR women) | 23 records |
| CINAHL Complete | 18/05/2021 | (Twitter OR tweet OR tweeting OR tweets OR retweet* OR tweep*) in Title  AND  (age OR gender OR demograph* OR minor* OR baby boomer* OR generation X OR generation Y OR generation Z OR gen X OR gen Y OR gen Z OR millennial* OR adult* OR teen* OR youth* OR adolescen* OR juvenile* OR young OR youngster* OR aged OR aging OR ageing OR elder* OR old OR retired OR older* OR geriatr* OR gerontolog* OR senior* OR senescen* OR retiree* OR sexagenarian* OR septuagenarian* OR octagenarian* OR nonagenarian* OR centenarian* OR supercentenarian* OR veteran* OR pensioner* OR male* OR female* OR men OR women OR sex) in Title | 65 records |
| Embase <1980 to 2021 Week 19> | 18/05/2021 | 1  (Twitter or tweet or tweeting or tweets or retweet* or tweep*).ti,ab. (5718) 2  demography/ (279727) 3  demograph*.ti,ab. (586081) 4  age distribution/ (147433) 5  exp groups by age/ (11355955) 6  "minor (person)"/ (706) 7  (minor* or baby boomer* or generation X or generation Y or generation Z or gen X or gen Y or gen Z or millennial* or adult*).ti,ab. (2068747) 8  (teen* or youth* or adolescen* or juvenile* or (young adj2 (adult* or person* or individual* or people* or population* or man or men or wom#n)) or youngster*).ti,ab. (749837) 9  (aged or aging or ageing or elder* or old or retired or older* or geriatr* or gerontolog* or senior* or senescen* or retiree* or sexagenarian* or septuagenarian* or octagenarian* or nonagenarian* or centenarian* or supercentenarian* or veteran* or pensioner*).ti,ab. (3205034) 10  age.ti,ab. (3738110) 11  sex ratio/ (73055) 12  exp "gender and sex"/ (1024490) 13  gender.ti,ab. (545242) 14  (male* and female*).ti,ab. (747966) 15  (men and women).ti,ab. (412683) 16  sex.ti,ab. (733711) 17  or/2-16 (14116212) 18  1 and 17 (2396) 19  exp algorithm/ (430675) 20  exp machine learning/ (249869) 21  exp algorithm/ (430675) 22  Neural Network*.ti,ab. (71079) 23  deep learning.ti,ab. (18237) 24  back propagation.ti,ab. (2883) 25  regression tree.ti,ab. (3502) 26  class prior.ti,ab. (103) 27  Conditional Random Field*.ti,ab. (632) 28  Decision Table.ti,ab. (137) 29  discriminating.ti,ab. (38906) 30  Decision Stump.ti,ab. (16) 31  Elastic Net.ti,ab. (1774) 32  Factor Graph Model*.ti,ab. (7) 33  Gaussian Mixture Model*.ti,ab. (1461) 34  Gaussian Process.ti,ab. (1315) 35  Higher Order Singular Value Decomposition.ti,ab. (53) 36  Instance-based Learning.ti,ab. (45) 37  J48.ti,ab. (295) 38  JRip.ti,ab. (29) 39  machine learning.ti,ab. (46762) 40  algorithms.ti,ab. (129440) 41  automati*.ti,ab. (168866) 42  Multi-task Learning.ti,ab. (340) 43  Non-negative Matrix Factorization.ti,ab. (1039) 44  Non-negative Tensor Factorization.ti,ab. (18) 45  Perceptron.ti,ab. (3042) 46  Random Sample Consensus.ti,ab. (141) 47  Rule Based.ti,ab. (4399) 48  Radial Basis Function Network.ti,ab. (210) 49  learning machine.ti,ab. (1426) 50  Reptree.ti,ab. (30) 51  Skip Gram.ti,ab. (48) 52  Support Vector Machine.ti,ab. (16434) 53  XGBoost.ti,ab. (861) 54  Stochastic gradient descent.ti,ab. (340) 55  Artificial intelligence.ti,ab. (14435) 56  Language Processing.ti,ab. (8475) 57  bag of words.ti,ab. (353) 58  text mining.ti,ab. (2787) 59  gated recurrent unit.ti,ab. (165) 60  word2vec.ti,ab. (170) 61  Determin*.ti. (351544) 62  Infer*.ti. (74214) 63  Ascertain.ti. (294) 64  Establish*.ti. (55917) 65  Predict*.ti. (507527) 66  Classify.ti,ab. (63529) 67  Classification.ti,ab. (453687) 68  Classifier.ti,ab. (27837) 69  intrinsic bias.ti,ab. (103) 70  facial recognition.ti,ab. (985) 71  extract*.ti. (182221) 72  identify.ti. (34537) 73  identifying.ti. (36813) 74  identified.ti. (28390) 75  or/20-74 (2373702) 76  18 and 75 (281) 77  twitter user*.ti,ab. (397) 78  75 and 77 (113) 79  76 or 78 (358) 80  remove duplicates from 79 (353) | 353 records |
| Google Scholar | 26/06/2020 | twitter age gender demographic | 767,000. Sifted until 100 non-relevant records |
| [IEEE/IET Electronic Library (IEEE Xplore)](http://libproxy.york.ac.uk/login?url=https://ieeexplore.ieee.org/Xplore/home.jsp) | 26/06/2020 | (Twitter OR tweet OR tweeting OR tweets OR retweet* OR tweep*) in Title  AND  (age OR gender OR demograph*) in Title | 25 records |
| LISTA | 18/05/2021 | (Twitter OR tweet OR tweeting OR tweets OR retweet* OR tweep*) in Title  AND  (age OR gender OR demograph* OR minor* OR baby boomer* OR generation X OR generation Y OR generation Z OR gen X OR gen Y OR gen Z OR millennial* OR adult* OR teen* OR youth* OR adolescen* OR juvenile* OR young OR youngster* OR aged OR aging OR ageing OR elder* OR old OR retired OR older* OR geriatr* OR gerontolog* OR senior* OR senescen* OR retiree* OR sexagenarian* OR septuagenarian* OR octagenarian* OR nonagenarian* OR centenarian* OR supercentenarian* OR veteran* OR pensioner* OR male* OR female* OR men OR women OR sex) in Title | 55 records |
| Proquest Dissertations & Theses: UK & Ireland | 18/05/2021 | (Twitter OR tweet OR tweeting OR tweets OR retweet* OR tweep*) in Document title  AND  (age OR gender OR demograph* OR minor* OR baby boomer* OR generation X OR generation Y OR generation Z OR gen X OR gen Y OR gen Z OR millennial* OR adult* OR teen* OR youth* OR adolescen* OR juvenile* OR young OR youngster* OR aged OR aging OR ageing OR elder* OR old OR retired OR older* OR geriatr* OR gerontolog* OR senior* OR senescen* OR retiree* OR sexagenarian* OR septuagenarian* OR octagenarian* OR nonagenarian* OR centenarian* OR supercentenarian* OR veteran* OR pensioner* OR male* OR female* OR men OR women OR sex) in Document title | 69 records |
| Ovid MEDLINE(R) and Epub Ahead of Print, In-Process & Other Non-Indexed Citations and Daily <1946 to May 17, 2021> | 18/05/2021 | 1  (Twitter or tweet or tweeting or tweets or retweet* or tweep*).ti,ab. (4310) 2  demography/ (61705) 3  demograph*.ti,ab. (352939) 4  age distribution/ (67431) 5  exp age groups/ (9394176) 6  minors/ (2642) 7  (minor* or baby boomer* or generation X or generation Y or generation Z or gen X or gen Y or gen Z or millennial* or adult*).ti,ab. (1619423) 8  (teen* or youth* or adolescen* or juvenile* or (young adj2 (adult* or person* or individual* or people* or population* or man or men or wom#n)) or youngster*).ti,ab. (602179) 9  (aged or aging or ageing or elder* or old or retired or older* or geriatr* or gerontolog* or senior* or senescen* or retiree* or sexagenarian* or septuagenarian* or octagenarian* or nonagenarian* or centenarian* or supercentenarian* or veteran* or pensioner*).ti,ab. (2373166) 10  age.ti,ab. (2403830) 11  exp sex distribution/ (65276) 12  exp Gender Identity/ (20455) 13  gender.ti,ab. (338094) 14  (male* and female*).ti,ab. (502290) 15  (men and women).ti,ab. (301108) 16  sex.ti,ab. (546438) 17  or/2-16 (11685101) 18  1 and 17 (1122) 19  classification/ (10421) 20  exp algorithms/ (347247) 21  Data Mining/ (9160) 22  Neural Network*.ti,ab. (58468) 23  deep learning.ti,ab. (15501) 24  back propagation.ti,ab. (2303) 25  regression tree.ti,ab. (2638) 26  class prior.ti,ab. (50) 27  Conditional Random Field*.ti,ab. (661) 28  Decision Table.ti,ab. (90) 29  discriminating.ti,ab. (31208) 30  Decision Stump.ti,ab. (10) 31  Elastic Net.ti,ab. (1181) 32  Factor Graph Model*.ti,ab. (7) 33  Gaussian Mixture Model*.ti,ab. (1248) 34  Gaussian Process.ti,ab. (1324) 35  Higher Order Singular Value Decomposition.ti,ab. (47) 36  Instance-based Learning.ti,ab. (43) 37  J48.ti,ab. (200) 38  JRip.ti,ab. (23) 39  machine learning.ti,ab. (38527) 40  algorithms.ti,ab. (104671) 41  automati*.ti,ab. (132916) 42  Multi-task Learning.ti,ab. (309) 43  Non-negative Matrix Factorization.ti,ab. (789) 44  Non-negative Tensor Factorization.ti,ab. (16) 45  Perceptron.ti,ab. (2613) 46  Random Sample Consensus.ti,ab. (117) 47  Rule Based.ti,ab. (3895) 48  Radial Basis Function Network.ti,ab. (175) 49  learning machine.ti,ab. (912) 50  Reptree.ti,ab. (27) 51  Skip Gram.ti,ab. (50) 52  Support Vector Machine.ti,ab. (13313) 53  XGBoost.ti,ab. (689) 54  Stochastic gradient descent.ti,ab. (260) 55  Artificial intelligence.ti,ab. (11535) 56  Language Processing.ti,ab. (7111) 57  bag of words.ti,ab. (324) 58  text mining.ti,ab. (2569) 59  gated recurrent unit.ti,ab. (140) 60  word2vec.ti,ab. (160) 61  Determin*.ti. (364254) 62  Infer*.ti. (66596) 63  Ascertain.ti. (264) 64  Establish*.ti. (47929) 65  Predict*.ti. (357533) 66  Classify.ti,ab. (47150) 67  Classification.ti,ab. (340482) 68  Classifier.ti,ab. (20726) 69  intrinsic bias.ti,ab. (86) 70  facial recognition.ti,ab. (755) 71  extract*.ti. (155990) 72  identify.ti. (24499) 73  identifying.ti. (28622) 74  identified.ti. (23424) 75  or/19-74 (1900871) 76  18 and 75 (175) 77  twitter user*.ti,ab. (353) 78  75 and 77 (109) 79  76 or 78 (253) | 253 records |
| APA PsycInfo <1987 to May Week 2 2021> | 18/05/2021 | 1  (Twitter or tweet or tweeting or tweets or retweet* or tweep*).ti,ab. (3362) 2  demograph*.ti,ab. (122659) 3  (minor* or baby boomer* or generation X or generation Y or generation Z or gen X or gen Y or gen Z or millennial* or adult*).ti,ab. (476915) 4  (teen* or youth* or adolescen* or juvenile* or (young adj2 (adult* or person* or individual* or people* or population* or man or men or wom#n)) or youngster*).ti,ab. (358974) 5  (aged or aging or ageing or elder* or old or retired or older* or geriatr* or gerontolog* or senior* or senescen* or retiree* or sexagenarian* or septuagenarian* or octagenarian* or nonagenarian* or centenarian* or supercentenarian* or veteran* or pensioner*).ti,ab. (590117) 6  age.ti,ab. (480959) 7  gender.ti,ab. (211626) 8  (male* and female*).ti,ab. (157447) 9  (men and women).ti,ab. (96265) 10  sex.ti,ab. (143406) 11  or/2-10 (1552274) 12  1 and 11 (828) 13  Neural Network*.ti,ab. (16828) 14  deep learning.ti,ab. (1500) 15  back propagation.ti,ab. (476) 16  regression tree.ti,ab. (330) 17  class prior.ti,ab. (20) 18  Conditional Random Field*.ti,ab. (84) 19  Decision Table.ti,ab. (39) 20  discriminating.ti,ab. (6938) 21  Decision Stump.ti,ab. (0) 22  Elastic Net.ti,ab. (175) 23  Factor Graph Model*.ti,ab. (1) 24  Gaussian Mixture Model*.ti,ab. (255) 25  Gaussian Process.ti,ab. (256) 26  Higher Order Singular Value Decomposition.ti,ab. (4) 27  Instance-based Learning.ti,ab. (64) 28  J48.ti,ab. (26) 29  JRip.ti,ab. (7) 30  machine learning.ti,ab. (6235) 31  algorithms.ti,ab. (14019) 32  automati*.ti,ab. (33432) 33  Multi-task Learning.ti,ab. (76) 34  Non-negative Matrix Factorization.ti,ab. (149) 35  Non-negative Tensor Factorization.ti,ab. (3) 36  Perceptron.ti,ab. (509) 37  Random Sample Consensus.ti,ab. (7) 38  Rule Based.ti,ab. (1894) 39  Radial Basis Function Network.ti,ab. (35) 40  learning machine.ti,ab. (473) 41  Reptree.ti,ab. (3) 42  Skip Gram.ti,ab. (20) 43  Support Vector Machine.ti,ab. (1786) 44  XGBoost.ti,ab. (26) 45  Stochastic gradient descent.ti,ab. (57) 46  Artificial intelligence.ti,ab. (3855) 47  Language Processing.ti,ab. (5575) 48  bag of words.ti,ab. (103) 49  text mining.ti,ab. (548) 50  gated recurrent unit.ti,ab. (16) 51  word2vec.ti,ab. (28) 52  Determin*.ti. (21937) 53  Infer*.ti. (9275) 54  Ascertain.ti. (32) 55  Establish*.ti. (4278) 56  Predict*.ti. (78877) 57  Classify.ti,ab. (9485) 58  Classification.ti,ab. (52361) 59  Classifier.ti,ab. (3470) 60  intrinsic bias.ti,ab. (20) 61  facial recognition.ti,ab. (658) 62  extract*.ti. (2363) 63  identify.ti. (2814) 64  identifying.ti. (7663) 65  identified.ti. (2351) 66  or/13-65 (257359) 67  12 and 66 (85) 68  twitter users.ti,ab. (260) 69  66 and 68 (43) 70  67 or 69 (115) | 115 records |
| Science Citation Index (SCI), Social Science Citation Index (SSCI), Conference Proceedings Citation Index – Science, Conference Proceedings Citation Index – Social Science and Humanities, Emerging Sources Citation Index (ESCI) --2015-present | 18/05/2021 | (TI=(determin* OR infer* OR ascertain OR establish* OR predict* OR extract* OR identify OR identifying OR identified) OR TS=(classify OR classification OR Classifier OR intrinsic bias OR facial recognition OR algorithm* OR machine learning OR Neural Network* OR deep learning OR back propagation OR regression tree OR class prior OR Conditional Random Field* OR Decision Table OR discriminating OR Decision Stump OR Elastic Net OR Factor Graph Model* OR Gaussian Mixture Model* OR Gaussian Process OR Higher Order Singular Value Decomposition OR Instance-based Learning OR J48 OR JRip OR automati* OR Multi-task Learning OR Non-negative Matrix Factorization OR Non-negative Tensor Factorization OR Perceptron OR Random Sample Consensus OR Rule Based OR Radial Basis Function Network OR learning machine OR Reptree OR Skip Gram OR Support Vector Machine OR XGBoost OR Stochastic gradient descent OR Artificial intelligence OR Language Processing OR bag of words OR text mining OR gated recurrent unit OR word2vec)) AND TS=(Twitter OR tweet OR tweeting OR tweets OR retweet* OR tweep*) AND TI=(age OR gender OR demograph* OR minor* OR baby boomer* OR generation X OR generation Y OR generation Z OR gen X OR gen Y OR gen Z OR millennial* OR adult* OR teen* OR youth* OR adolescen* OR juvenile* OR young OR youngster* OR aged OR aging OR ageing OR elder* OR old OR retired OR older* OR geriatr* OR gerontolog* OR senior* OR senescen* OR retiree* OR sexagenarian* OR septuagenarian* OR octagenarian* OR nonagenarian* OR centenarian* OR supercentenarian* OR veteran* OR pensioner* OR male* OR female* OR men OR women OR sex) | 197 records |
| Zetoc | 26/06/2020 | Multiple searches of the title field were carried out:  Twitter AND gender (16 hits)  Twitter AND age (25 hits)  Twitter AND Demographics (5 hits)  Twitter AND Demographic (15 hits) | 61 records hits (including duplicates) |
